# Linear epitopes of SARS-CoV-2 spike protein elicit neutralizing antibodies in COVID-19 patients

**DOI:** 10.1101/2020.06.07.20125096

**Authors:** Yang Li, Dan-yun Lai, Hai-nan Zhang, He-wei Jiang, Xiao-long Tian, Ming-liang Ma, Huan Qi, Qing-feng Meng, Shu-juan Guo, Yan-ling Wu, Wei Wang, Xiao Yang, Da-wei Shi, Jun-biao Dai, Tian-lei Ying, Jie Zhou, Sheng-ce Tao

**Author notes:** **Corresponding:** (S.-C. Tao); (J. Zhou); (T.-L. Ying). These authors contributed equally to this work (Yang Li, Dan-yun Lai, Hai-nan Zhang, He-wei Jiang, Xiao-long Tian).

## Abstract

SARS-CoV-2 outbreak is a world-wide pandemic. The Spike protein plays central role in cell entry of the virus, and triggers significant immuno-response. Our understanding of the immune-response against S protein is still very limited. Herein, we constructed a peptide microarray and analyzed 55 convalescent sera, three areas with rich linear epitopes were identified. Potent neutralizing antibodies enriched from sera by 3 peptides, which do not belong to RBD were revealed.

---

COVID-19 is caused by SARS-CoV-2^1,2^. By June 8th, 2020, globally, 7,007,948 diagnosed cases, 402,709 deaths were reported (https://coronavirus.jhu.edu/map.html)^3^.

High titer of Spike protein (S protein) specific antibodies is in the blood of COVID-19 patients, especially IgG for both SARS-CoV^4^ and SARS-CoV-2^5,6^. Because of the central role that S protein plays in the entry of virus to the host cell, S1 and more specific, RBD (Receptor Binding Domain) is the most-focused target for the development of COVID-19 therapeutic antibodies^7,8^ and vaccines^9^. It is known that besides RBD, other areas/ epitopes of S protein may also elicit neutralization antibodies^10^. However, antibody responses to full length S protein at epitope resolution has not been investigated and the capability of linear epitopes to elicit neutralizing antibody is still not explored.

To precisely decipher the B cell linear epitopes of S protein, we constructed a peptide microarray. A total of 211 peptides (**Extended Data Table 1**) were synthesized and conjugated to BSA. (**Extended Data Fig.1a-c)**. The conjugates along with control proteins were printed in triplicate, and with three dilutions. High reproducibility among triplicated spots or repeated arrays for serum profiling were achieved (**Extended Data Fig.1d-e)**. The peptides with variant concentrations may enable dynamical detection of antibody responses and indicate the antibodies against different epitopes may have different kinetic characteristics (**Extended Data Fig.1f, Extended Data Fig.2a)**. Moreover, inhibitory assay using free peptides verified the specificity of the signals generated against the peptides (**Extended Data Fig.2b)**.

Fifty-five sera from convalescent COVID-19 patients and 18 control sera (**Extended Data Table 2**) were screened on the peptide microarray for both IgG and IgM responses (**Fig. 1a** and **Extended Data Fig. 3)**. For IgG, COVID-19 patients were completely separated from controls, distinct and specific signals were shown for some peptides. In contrast, it was not distinct enough for IgM responses. We then focused on IgG for further analysis. Epitope map of S protein were generated based on the response frequency (**Fig.1b**).

**Fig. 1.**
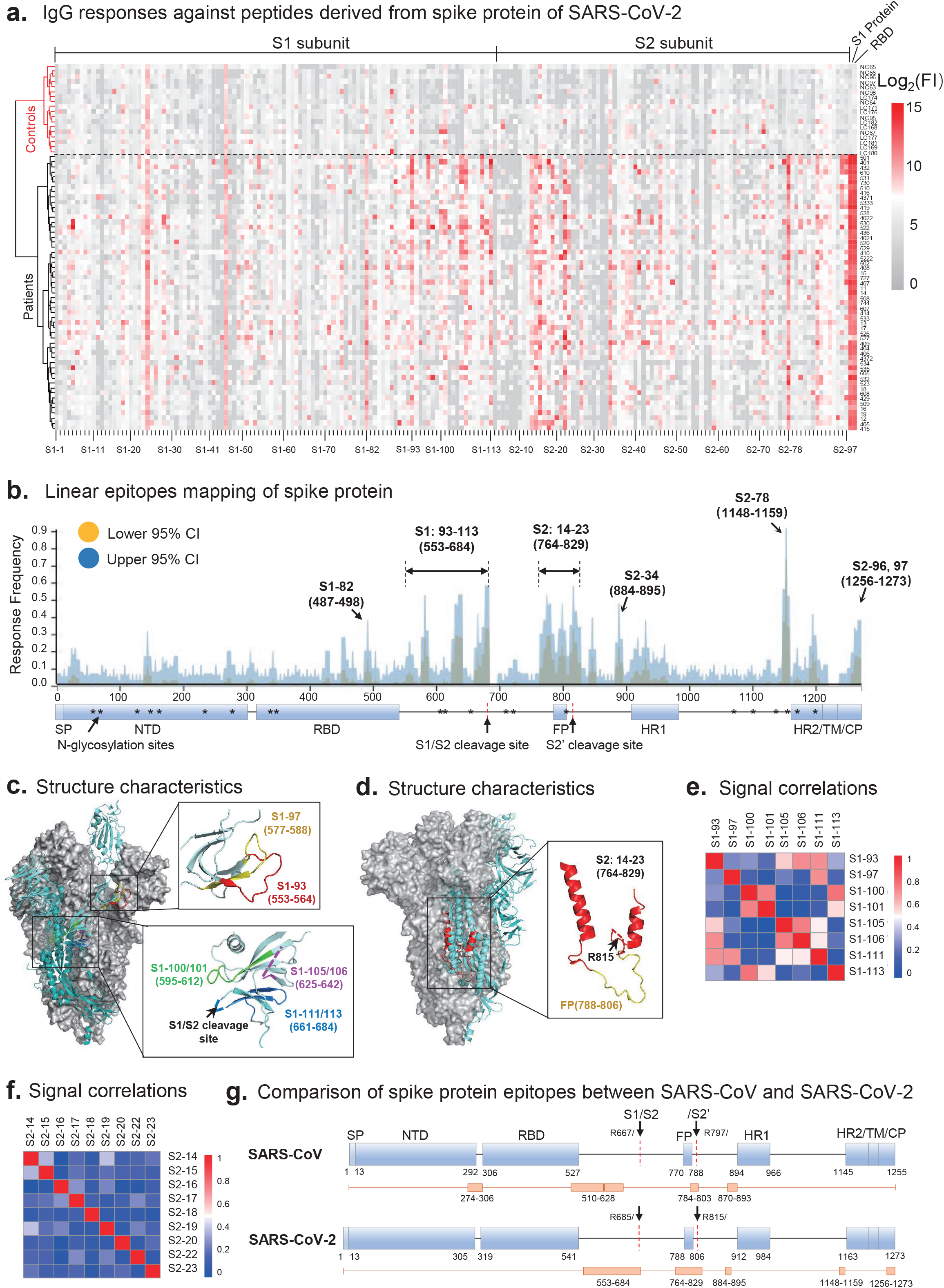
Linear epitope mapping of SARS-CoV-2 S protein specific antibodies by a peptide microarray, the IgG channel. **a**. Heatmap of IgG antibody responses of 55 sera from COVID-19 convalescent patients and controls (Healthy donors and Lung cancer patients). FI, fluorescent intensity. **b**. Epitope mapping according to the response frequency. CI, confidence interval. **c-d**. Detailed structural information of the epitopes of two hot areas on S protein (PDB: 6vyb). **e-f**, Correlations of the antibody responses among the peptides for the two hot areas. **g**. The B cell epitopes (labeled by orange rectangles) on the corresponding positions of S protein for both SARS-CoV^11^ and SARS-CoV-2.

Majorly, there are three hot epitope areas across S protein. The first is on CTD (C Terminal Domain) that immediately follows RBD, *i*.*e*. from S1-93 to S1-113. Interestingly, the identified epitopes, S1-93, 97, 100/101, 105/106, 111 and 113 locate predominantly at flexible loops (**Fig. 1c**). In addition, the signals of some epitopes had moderate correlations with others (**Fig. 1e**), and most of these epitopes were positively correlated with S1 (**Extended Data Fig. 4c-f**. The second hot area is from S2-14 to S2-23, including the FP (Fusion Peptide, aa788-806) region and the S2’cleave site (R815) (**Fig. 1d)**. In contrary to the first hot region, the antibody responses against epitopes of this region had poor correlations among each other (**Fig. 1f)**, possibly due to the capability of this region to generate continuous but competitive epitopes. Moreover, part of this area is shielded by other parts in trimeric S (**Fig. 1d)**, suggesting this part would be easily accessed by immune system after the depart of S2 from S1. The third hot area is S2-78 or aa1148-1159, connecting HR1 (Heptad Repeat 1) and HR2 (Heptad Repeat 2) on S2 subunit. IgG antibodies against this epitope can be detected in about 90% COVID-19 patients, indicating it is an extremely dominant epitope. Except for these three areas, S2-34 (aa884-895) and S2-96/97 (1256-1273) also elicited antibodies in some patients. Overall, the epitope pattern of SARS-CoV-2 S protein is similar to that of SARS-CoV^11^ (**Fig. 1g)**.

RBD can elicit high titer of antibodies and highly correlates with that of S1 protein ^6^ (**Extended Data Fig.4a**), suggesting RBD is a dominant region. A peptide, S1-82, locates exactly on the surface of RBM (Receptor Binding Motif) (**Extended Data Fig 5a**), was identified as an epitope. However, further analysis demonstrated that the epitope had a poor specificity (**Fig. 2a, Extended Data Fig 5a**), probably due to the sequence similarity (**Extended Data Fig.5c-g**). Thus the validity of epitope S1-82 may need further investigation. Besides S1-82, no significant binding was observed for the rest of peptides locate at RBD, suggesting that conformational epitopes are dominant for RBD.

**Fig. 2.**
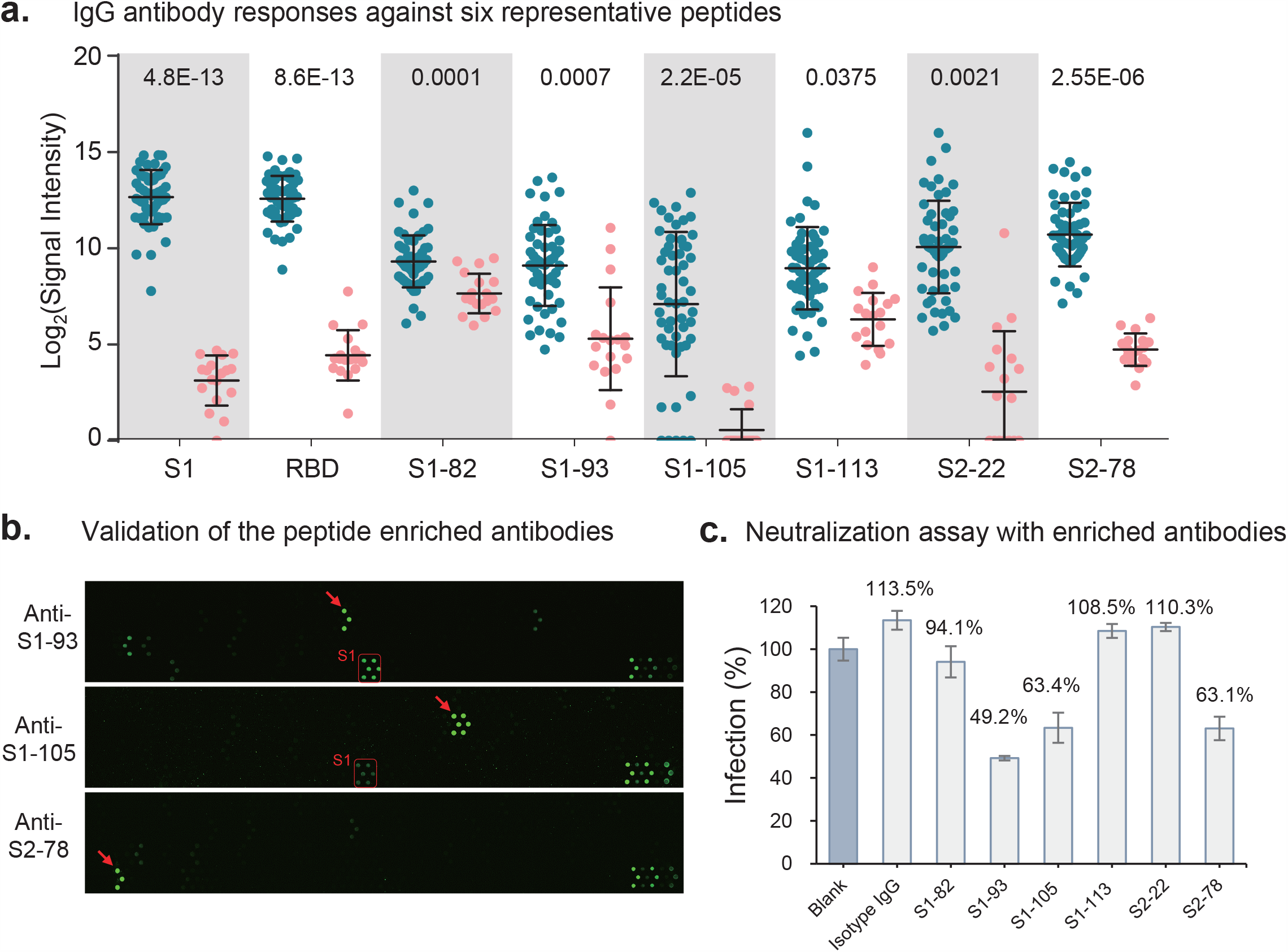
Evaluation of neutralizing activities of epitope-specific antibodies. **a**. IgG responses against six selected peptides of COVID-19 patients (green) and controls (pink). **b**. Peptide microarray results for the enriched epitope-specific antibodies. **c**. Neutralization assay with the epitope-specific antibodies. Relative infection rates for each sample to blank control are indicated.

Besides RBD, other areas/ epitopes of S protein may also elicit neutralization antibodies^10^. To explore this possibility, we chose 6 representative epitopes, *i*.*e*. S1-82, S1-93, S1-105, S1-113, S2-22 and S2-78 to test (**Fig. 2a**). Antibodies that specifically bind these epitopes were separately enriched from five sera. High specificity of these antibodies, except for S1-82, were demonstrated **(Fig. 2b, Extended Data Fig.6)**. Pseudotyped virus neutralization assay with the enriched antibodies was then performed. Because of the limited amount of the enriched epitope-specific antibodies, the assay was performed for only a single point with the highest antibody concentration that was applicable for each epitope-specific antibody. Surprisingly, the antibodies against three epitopes, *i*.*e*., S1-93, S1-105 and S2-78 exhibited potent neutralizing activity with a 51%, 35%, 35% virus infection inhibitory efficiency at 8.3, 10.4 and 21 µg/mL, respectively (**Fig. 2c**). While not for S1-82, S1-113 and S2-22 at 2.6, 7.6 and 21 µg/mL, respectively.

S1-93 locates at CTD of S1. The antibody against this epitope has neutralizing activity, which is consistent with a recent study^12^, antibody binds this epitope may affect the conformation change of S for ACE2 binding. S1-105 also belongs to the CTD but close to the S1/S2 cleavage site. The antibodies may block effective protease cleavage on this site, which is critical for the entry of the virus. S2-78 locates adjacent to HR2, antibodies bind to this epitope may interfere the formation of 6-HB (Helical Bundle), an essential structure for cell membrane fusion^13^.

Potent neutralizing antibodies could provide therapeutic and prophylactic reagents to fight against the COVID-19 pandemic^11^. However, it is risky to only focus on the RBD region, evolutionary pressure on this “hot” area may cause potential mutations in this region. This may by pass or defect the effectiveness of the RBD region centered therapeutic antibodies and vaccines in the developing pipeline^14^. Thus identification of other domains or epitopes that can elicit neutralizing antibodies is essential as well. Combination of the potent antigenicity of these peptides and neutralizing activity of the corresponding antibodies makes these epitopes potential candidates for both therapeutic antibodies and vaccine development.

There are some limitations in this study. Because of very limited amount of purified antibodies could be enriched from valuable serum samples, we cannot perform a full set of neutralization assay by serial dilution to obtain the exact IC50. The IC50 estimated based on present data was 5-20 µg/mL for the antibodies. However, monoclonal antibodies presumably have a better neutralization activity, so it is an option to acquire the reactive B cell clone and express recombinant monoclonal antibodies^2,7^. Because of the poor availability of the convalescent sera, we did not thoroughly examine all the peptides that may potentially raise neutralizing antibodies. The other epitopes raised antibodies may also have neutralizing activities, such as S1-97, which is very closed to S1-93, or the peptides derived from the FP region, such as S2-18,19,20, which worth further investigation.

## Data Availability

The peptide microarray data are deposited on Protein Microarray Database (http://www.proteinmicroarray.cn) under the accession number PMDE242. Additional data related to this paper may be requested from the authors.

## Figure Legends

**Extended Data Fig. 1.**
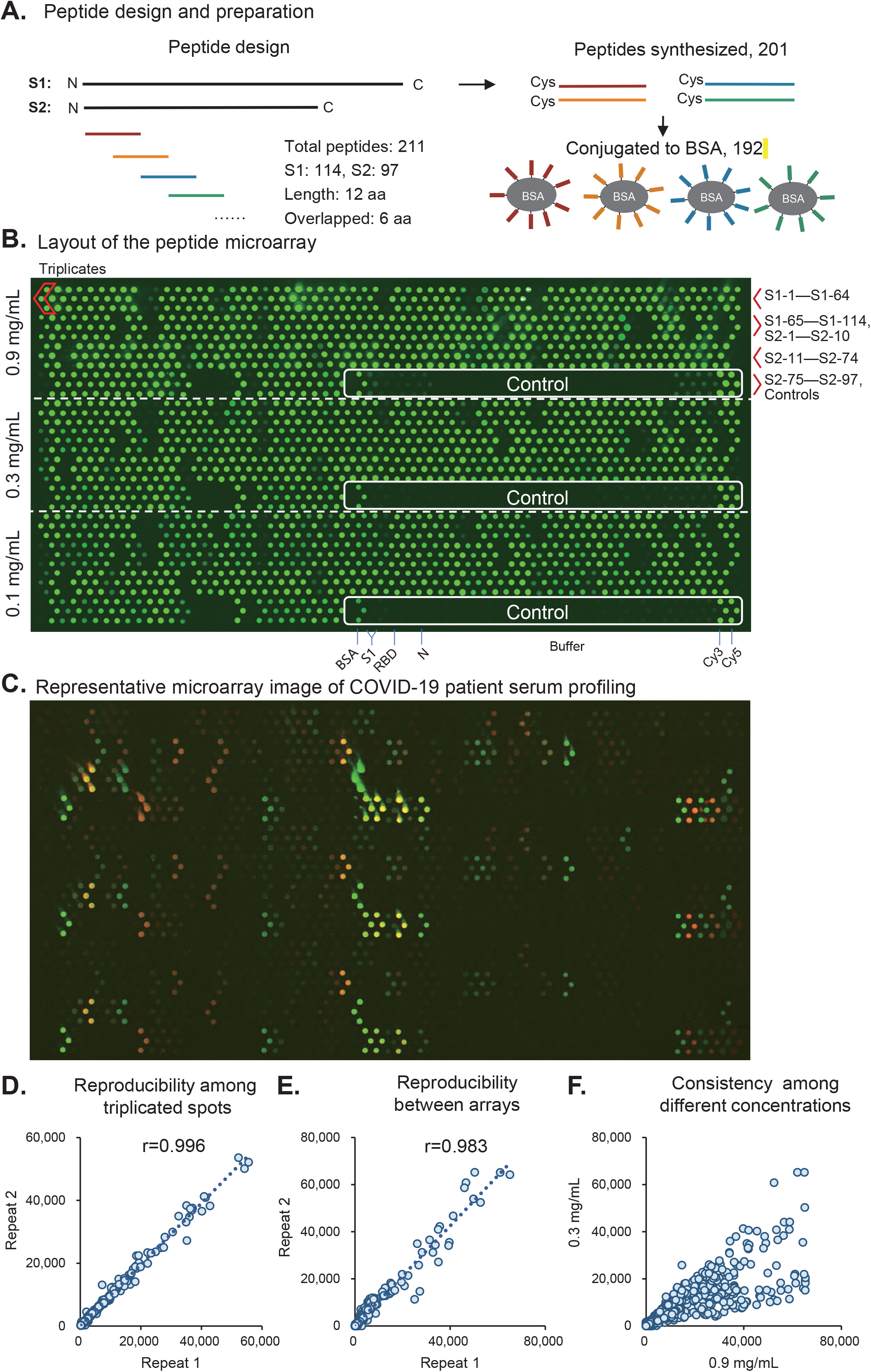
Peptide design and microarray fabrication. **a**. Peptide design and conjugation with BSA through the cysteine on the N terminal. The numbers 201 and 192 indicates the peptides that were successfully synthesized and conjugated, respectively. b. layout of the peptide microarray. The image was from anti-BSA antibody incubation. Peptides were sequentially printed and the peptides for each row are indicated on the right. **c**. A representative merged image of one COVID-19 serum. IgG response is indicated as green, while IgM is indicated as red. **d-f**. Correlations between repeated spots of the same protein on the same array, repeats arrays and peptide groups with different concentrations.

**Extended Data Fig. 2.**
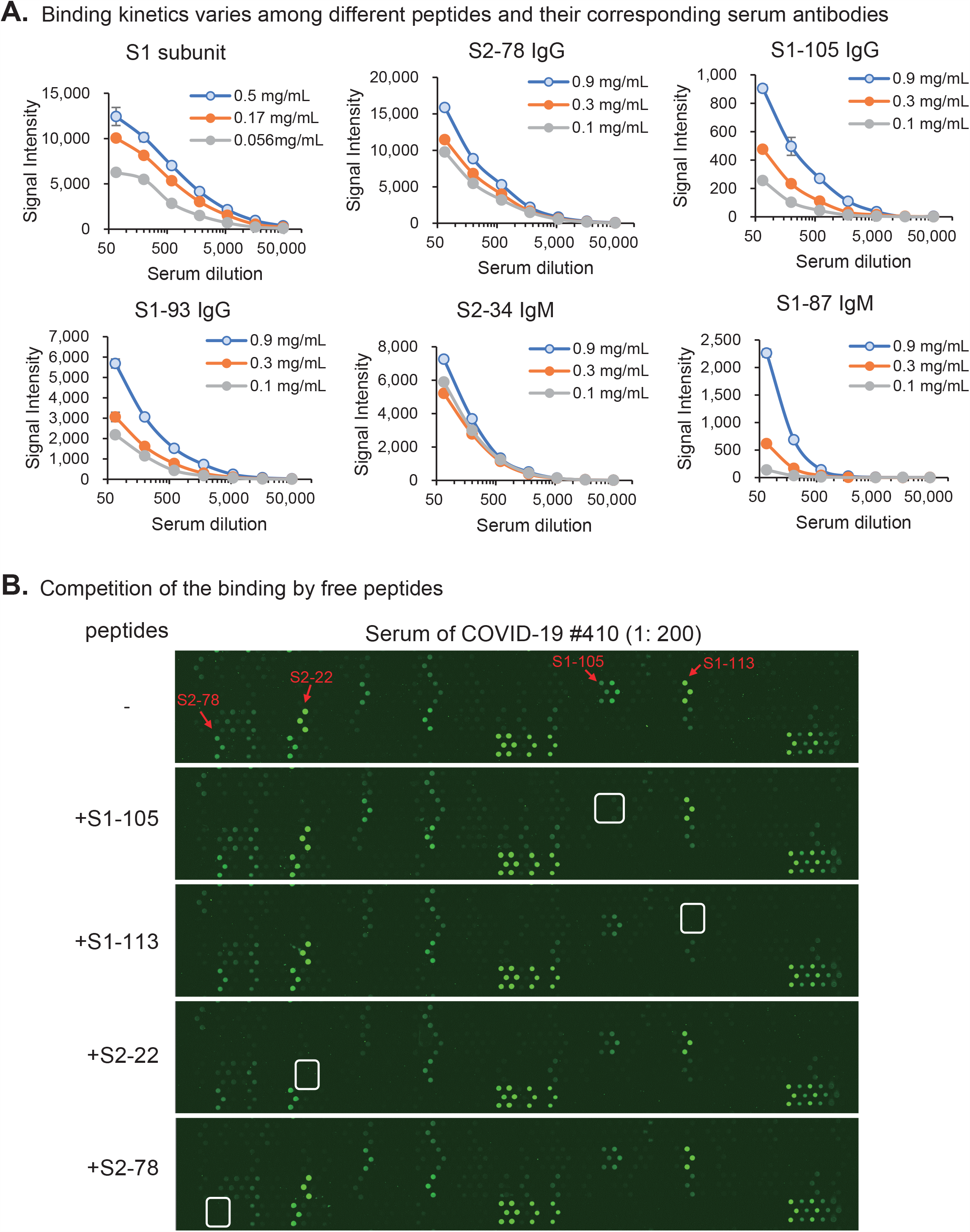
Detection of the SARS-CoV-2 specific antibody responses by using the peptide microarray. **a**. Dynamic change of signal intensities for some representative peptides and S1 protein in different concentrations. b. Microarray results of the competition assays with the addition of free peptides to the sera. The serum used and the dilution are labeled above. The peptides used for competition are labeled on the left and the red arrows and white rectangles indicate the position of the corresponding peptides.

**Extended Data Fig. 3.**
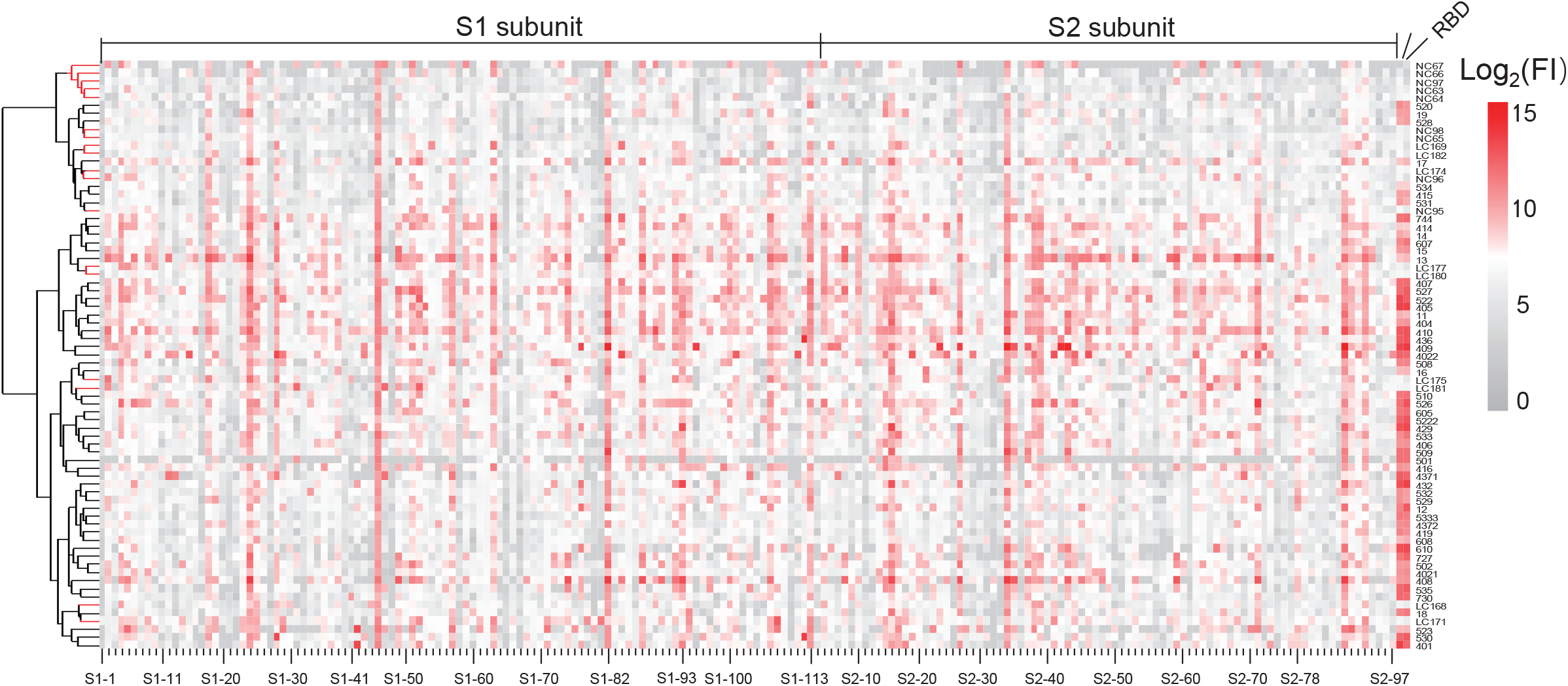
IgM responses against peptides derived from S protein. Heatmap of IgM antibody responses of 55 sera from COVID-19 convalescent patients and controls (Healthy donors and Lung cancer patients). Peptides that fully cover S protein were surveyed and S1 protein and RBD were included on the peptide microarray as controls. The peptides were sequentially arranged without clustering. FI, fluorescent intensity.

**Extended Data Fig. 4.**
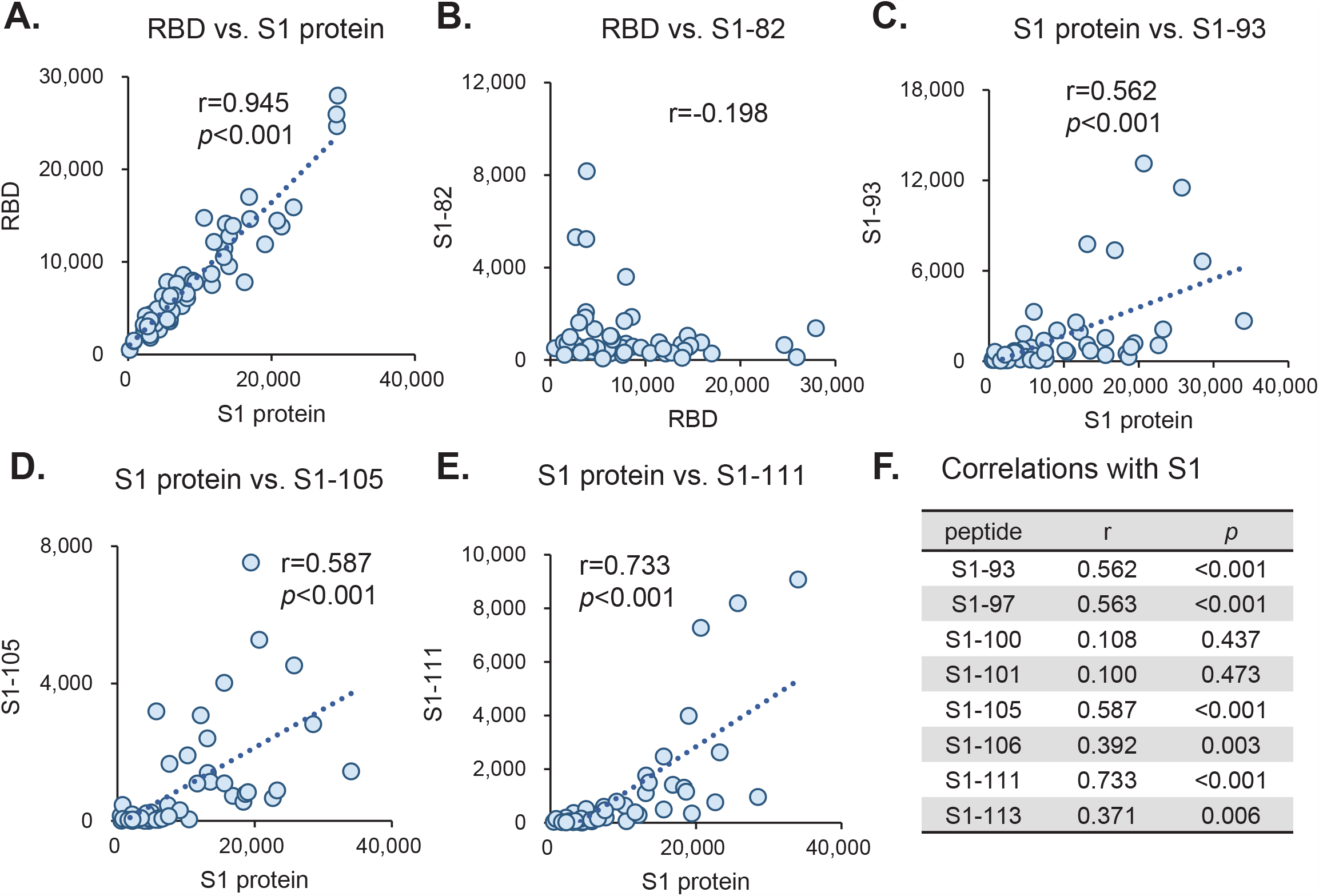
Correlation analysis among antibody responses against S1 subunit derived epitopes and S1 protein. **a-e**. Correlations of IgG responses between two peptides, proteins or peptide and proteins. **f**. Summary of the correlations of IgG responses between representative peptides and S1 protein.

**Extended Data Fig. 5.**
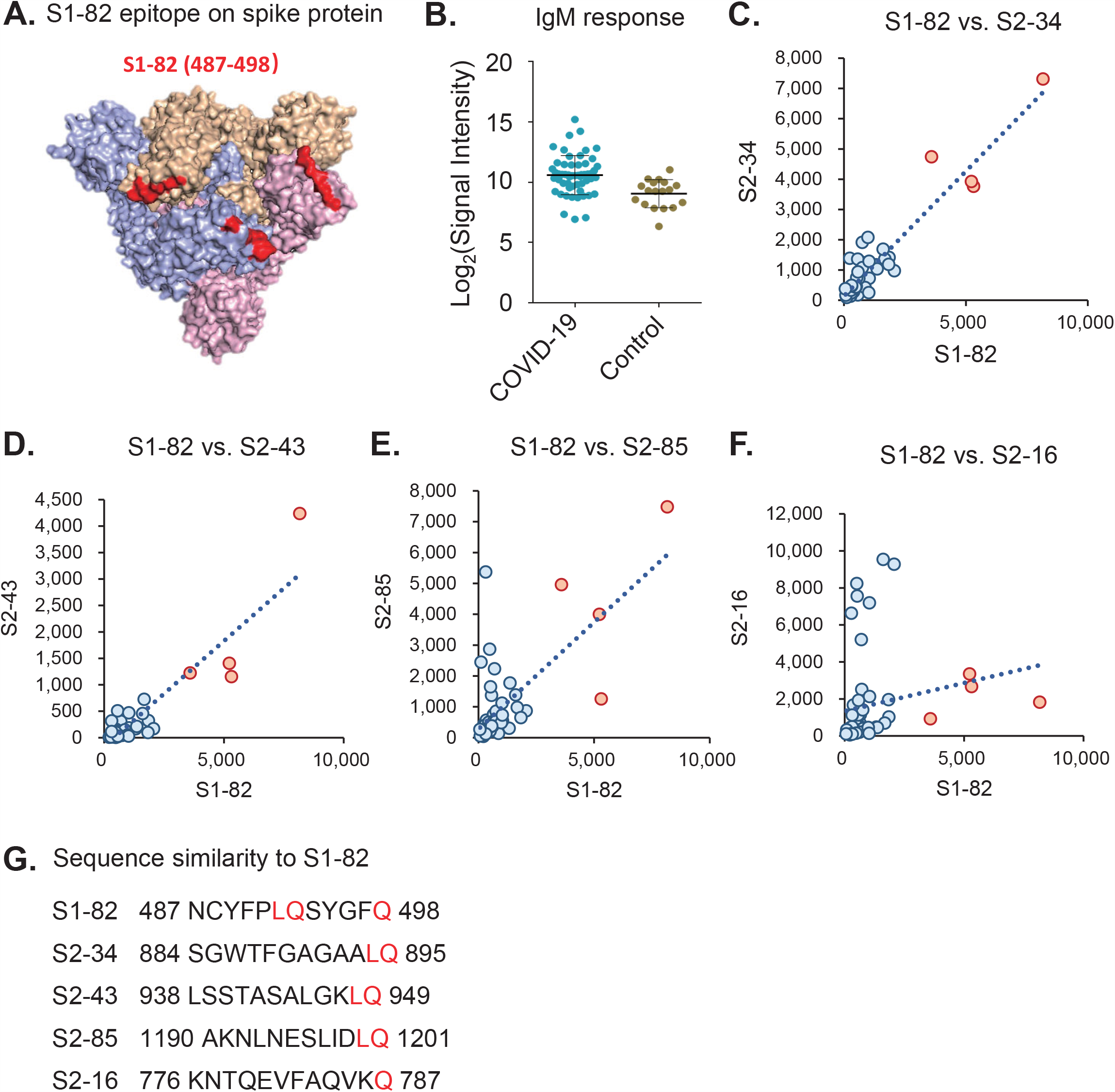
Cross-activity of anti-S2-82 antibodies. **a**. The structural position of S1-82 on S protein (PDB: 6xyb). **b**. IgM responses of S1-82 in COVID-19 patients (blue) and controls (yellow). c-f. correlations of IgG responses between S1-82 and other peptides. The red spots were samples with high IgG signals for S1-82. g. Sequence similarity among the indicated peptides with S1-82.

**Extended Data Fig. 6.**
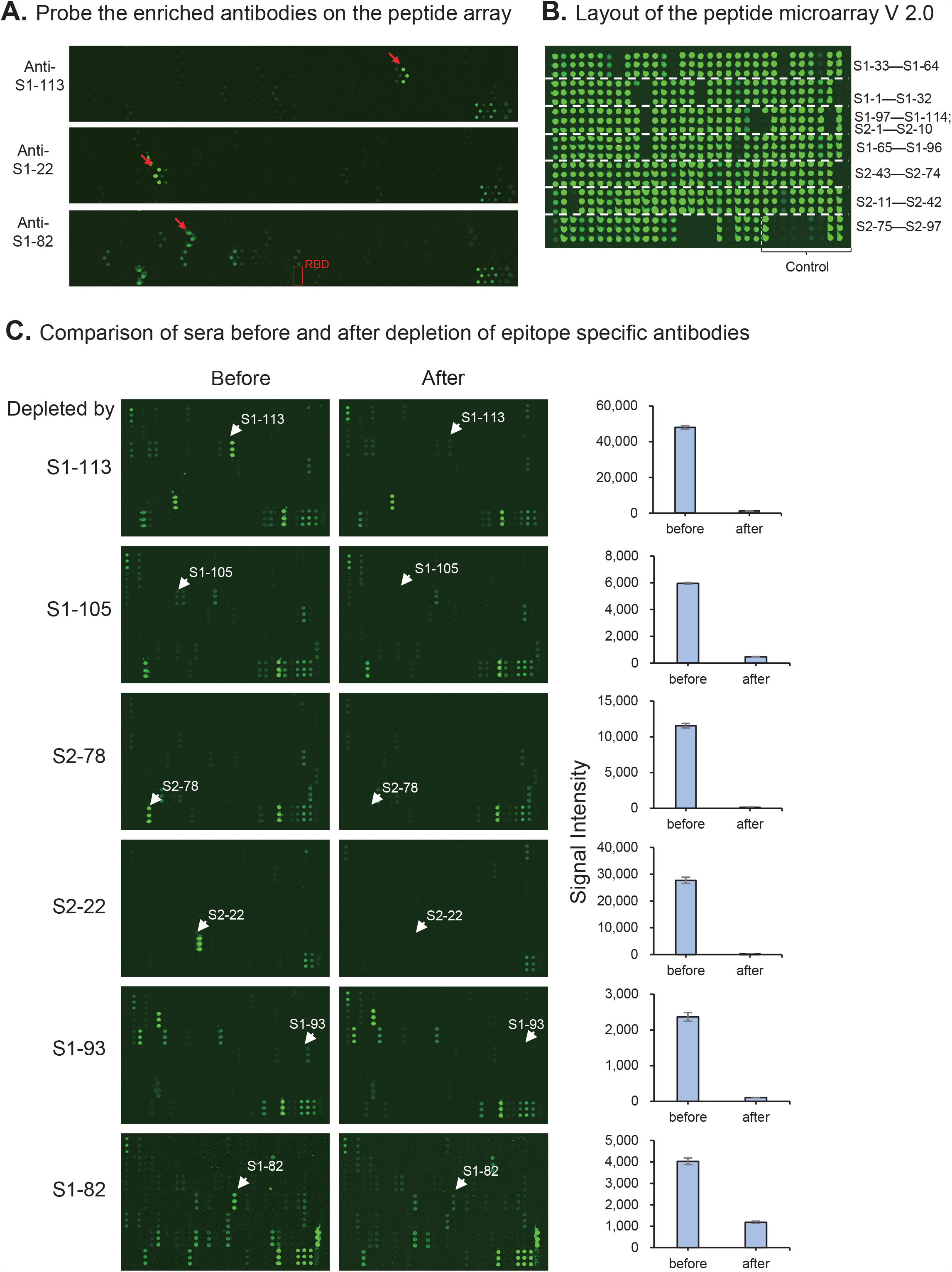
Epitope-specific antibody depletion from sera. **a**. Peptide microarray results for epitope-specific antibodies. Red arrows indicate the corresponding peptides. b. Layout of the new version of the peptide microarray with 0.3 mg/mL peptides printed. c. Representative images (left) and results (right) for comparison of sera between before and after depletion of epitope specific antibodies. The positions of the corresponding peptides labeled on the left that were used for depletion are indicated by arrows.

## Methods

### Peptide synthesis and conjugation with BSA

The N-terminal amidated peptides were synthesized by GL Biochem, Ltd. (Shanghai, China). Each peptide was individually conjugated with BSA using Sulfo-SMCC (Thermo Fisher Scientific, MA, USA) according to the manufacture’s instruction. Briefly, BSA was activated by Sulfo-SMCC in a molar ratio of 1: 30, followed by dialysis in PBS buffer. The peptide with cysteine was added in a w/w ratio of 1:1 and incubated for 2 h, followed by dialysis in PBS to remove free peptides. A few conjugates were randomly selected for examination by SDS-PAGE. For the conjugates of biotin-BSA-peptide, before conjugation, BSA was labelled with biotin by using NHS-LC-Biotin reagent (Thermo Fisher Scientific, MA, USA) with a molar ratio of 1: 5, and then activated by Sulfo-SMCC.

### Peptide microarray fabrication

The peptide-BSA conjugates as well as S1 protein, RBD protein and N protein of SARS-CoV-2, along with the negative (BSA) and positive controls (anti-Human IgG and IgM antibody), were printed in triplicate on PATH substrate slide (Grace Bio-Labs, Oregon, USA) to generate identical arrays in a 1 × 7 subarray format using Super Marathon printer (Arrayjet, UK). The microarrays were stored at -80°C until use.

### Patients and samples

The Institutional Ethics Review Committee of Foshan Fourth Hospital, Foshan, China approved this study and the written informed consent was obtained from each patient. COVID-19 patients were hospitalized and received treatment in Foshan Forth hospital during the period from 2020-1-25 to 2020-3-8 with variable stay time (**Extended data**

**Table 2**). Serum from each patient was collected on the day of hospital discharge when the standard criteria were met according to Diagnosis and Treatment Protocol for Novel Coronavirus Pneumonia (Trial Version 5), released by the National Health Commission & State Administration of Traditional Chinese Medicine. The basic criteria are the same with that in the Diagnosis and Treatment Protocol for Novel Coronavirus Pneumonia (Trial Version 7)^1^. Briefly, the key points of the discharge criteria are: 1) Body temperature is back to normal for more than three days; 2) Respiratory symptoms improve obviously; 3) Pulmonary imaging shows obvious absorption of inflammation; 4) Nuclei acid tests negative twice consecutively on respiratory tract samples such as sputum and nasopharyngeal swabs (sampling interval being at least 24 hours). From 2020-2-28, standard criteria of discharge were modified by adding one item that nuclei acid tests should be negative on anal swab sample. Sera of the control group from Lung cancer patients and healthy donors were collected from Ruijin Hospital, Shanghai, China. All the sera were inactivated at 56 °C for 30 min and stored at -80C until use.

### Microarray-based serum analysis

A 7-chamber rubber gasket was mounted onto each slide to create individual chambers for the 7 identical subarrays. The microarray was used for serum profiling as described previously with minor modifications^2^. Briefly, the arrays stored at -80°C were warmed to room temperature and then incubated in blocking buffer (3% BSA in 1×PBS buffer with 0.1% Tween 20) for 3 h. A total of 400 µL of diluted sera or antibodies was incubated with each subarray for 2 h. The sera were diluted at 1:200 for most samples and for competition experiment, free peptides were added at a concentration of 0.25 mg/mL. For the enriched antibodies, 0.1-0.5 µg antibodies were included in 400 µL incubation buffer. The arrays were washed with 1×PBST and bound antibodies were detected by incubating with Cy3-conjugated goat anti-human IgG and Alexa Fluor 647-conjugated donkey anti-human IgM (Jackson ImmunoResearch, PA, USA), which were diluted for 1: 1,000 in 1×PBST. The incubation was carried out at room temperature for 1 h. The microarrays were then washed with 1×PBST and dried by centrifugation at room temperature and scanned by LuxScan 10K-A (CapitalBio Corporation, Beijing, China) with the parameters set as 95% laser power/ PMT 550 and 95% laser power/ PMT 480 for IgM and IgG, respectively. The fluorescent intensity was extracted by GenePix Pro 6.0 software (Molecular Devices, CA, USA).

### Purification of epitope-specific antibodies

Depends on the availability,200-500 µL serum from COVID-19 convalescent patient was two-fold diluted in 1×PBS and then pre-incubated with streptavidin beads to eliminate non-specific binding. For each epitope, 100 μg peptides conjugated with biotin-BSA were coated to 100 μL streptavidin magnetic beads (Invitrogen, MA, USA) in 1×PBS buffer at room temperature for 1 h. The protein-coated streptavidin beads were washed 4 times in 1×PBS containing 0.1% BSA, and incubated with the pre-cleaned serum in 1×PBS at 4°C for 4 h. The streptavidin beads were then washed 3 times in 1×PBS containing 0.1% BSA, and eluted with 0.2 M glycine, 1 mM EGTA, pH 2.2. Finally, the antibodies were neutralized with 1M Tris-HCl, pH8.0. The concentration of the purified antibody was monitored by silver staining.

### Pseudotyped Virus Neutralization

The neutralization assay was performed as described^3^. Briefly, 293 T cells were co-transfected with expression vectors of pcDNA3.1-SARS-CoV-2-S (encoding SARS-CoV-2 S protein) and pNL4-3.luc.RE bearing the luciferase reporter-expressing HIV-1 backbone. The supernatants containing SARS-CoV-2 pseudotyped virus were collected 48 h post-transfection. Antibodies or isotype IgG control (Thermo Fisher Scientific, MA, USA) in DMEM supplemented with 10% fetal calf serum were incubated with pseudoviruses at 37C for 1 h and then the mixtures were added to monolayer Huh-7 cells (10^4^ per well in 96-well plates). Twelve h after infection, culture medium was refreshed and then the cells were incubated for an additional 48 h. The luciferase activity was calculated for the detection of relative light units using the Bright-Glo Luciferase Assay System (Promega, WI, USA). Huh-7 cells were subsequently lysed with 50 µl lysis reagent (Promega, WI, USA), and 30 µl of the lysates were transferred to 96-well Costar flat-bottom luminometer plates (Corning Costar, MA, USA) for the detection of relative light units using the Firefly Luciferase Assay Kit (Promega, WI, USA) and an Ultra 384 luminometer (Tecan, Switzerland).

### Data analysis and software

Signal Intensity was defined as the median of the foreground subtracted by the median of background for each spot and then averaged the triplicate spots for each peptide or protein. IgG and IgM data were analyzed separately. Pearson correlation coefficient between two proteins or indicators and the corresponding *p* value was calculated by SPSS software under the default parameters. Cluster analysis was performed by pheatmap package in R^4^. To calculate the response frequency of each epitope specific antibody, mean signal + 3*SD of the control sera were used to set the threshold. The epitope map was generated by ImmunomeBrowser issued by IEDB (Epitope Prediction and Analysis Tools)^5^. Visualization of the structural details were processed by Pymol (https://pymol.org/2/).

## Acknowledgements

This work was partially supported by the National Key Research and Development Program of China Grant (No. 2016YFA0500600), National Natural Science Foundation of China (No. 31970130, 31600672, 31670831, and 31370813), Shenzhen Key Laboratory of Synthetic Genomics (ZDSYS201802061806209), Shenzhen Science and Technology Program (KQTD20180413181837372), Guangdong Provincial Key Laboratory of Synthetic Genomics (2019B030301006) and Foshan Scientific and Technological Key Project for COVID-19 (NO:2020001000430).

## Author contributions

S-C. T. developed the conceptual ideas and designed the study. J. Z., W. W. and X. Y. collected the sera samples and provided key reagents. Y. L., D-Y. L., H-N. Z., X-L. T., H-W. J., M-L. M., H. Q., Q-F. M. and S-J. G. performed the experiments. T-L. Y and Y.L.-W provide critical suggestion on the neutralization assay. S-C.T. and Y. L. wrote the manuscript with suggestions from other authors. D-W. S. and J-B. D. provided key reagents or data analysis.

---

**Extended Data Table 1.**
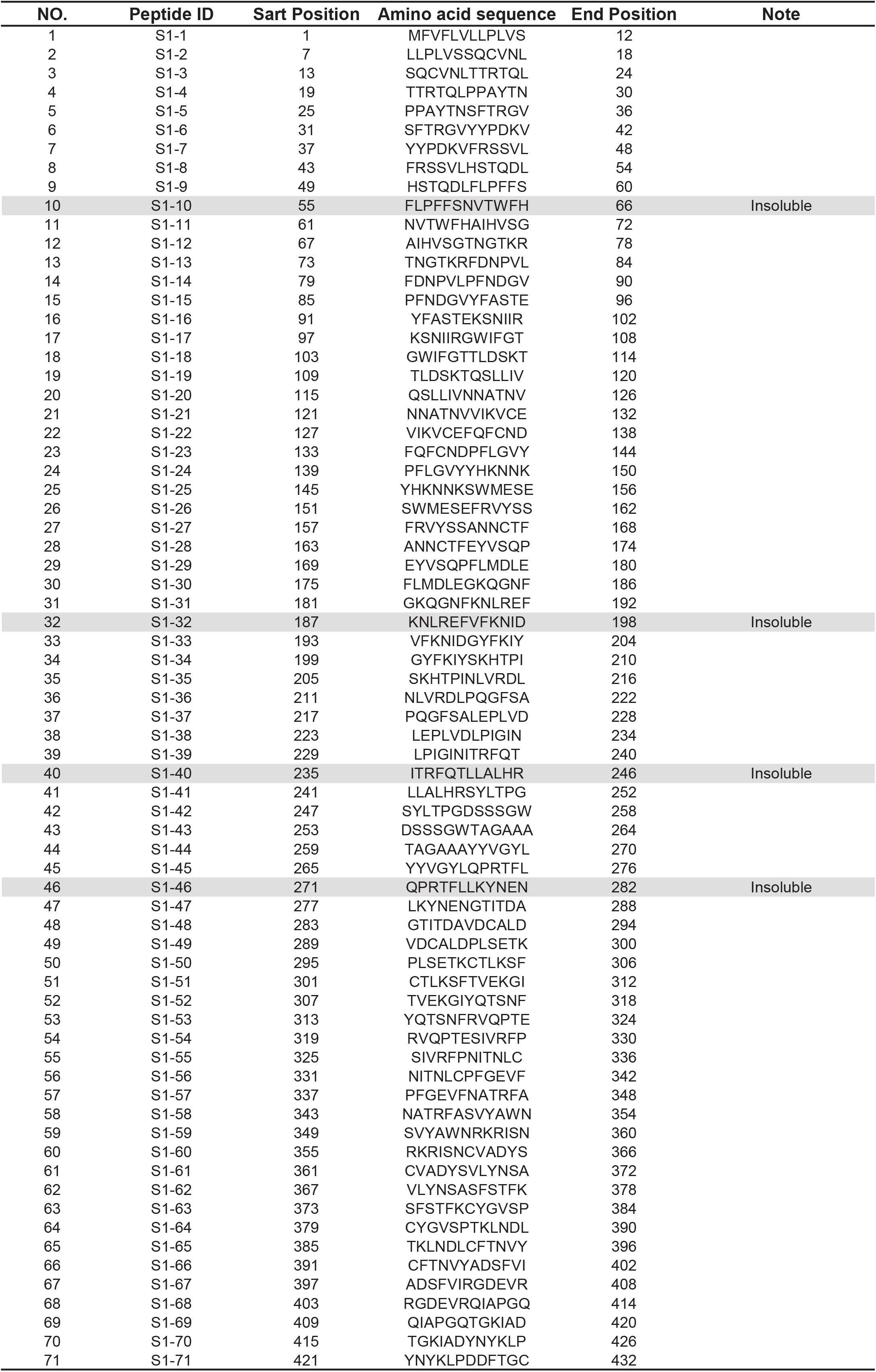
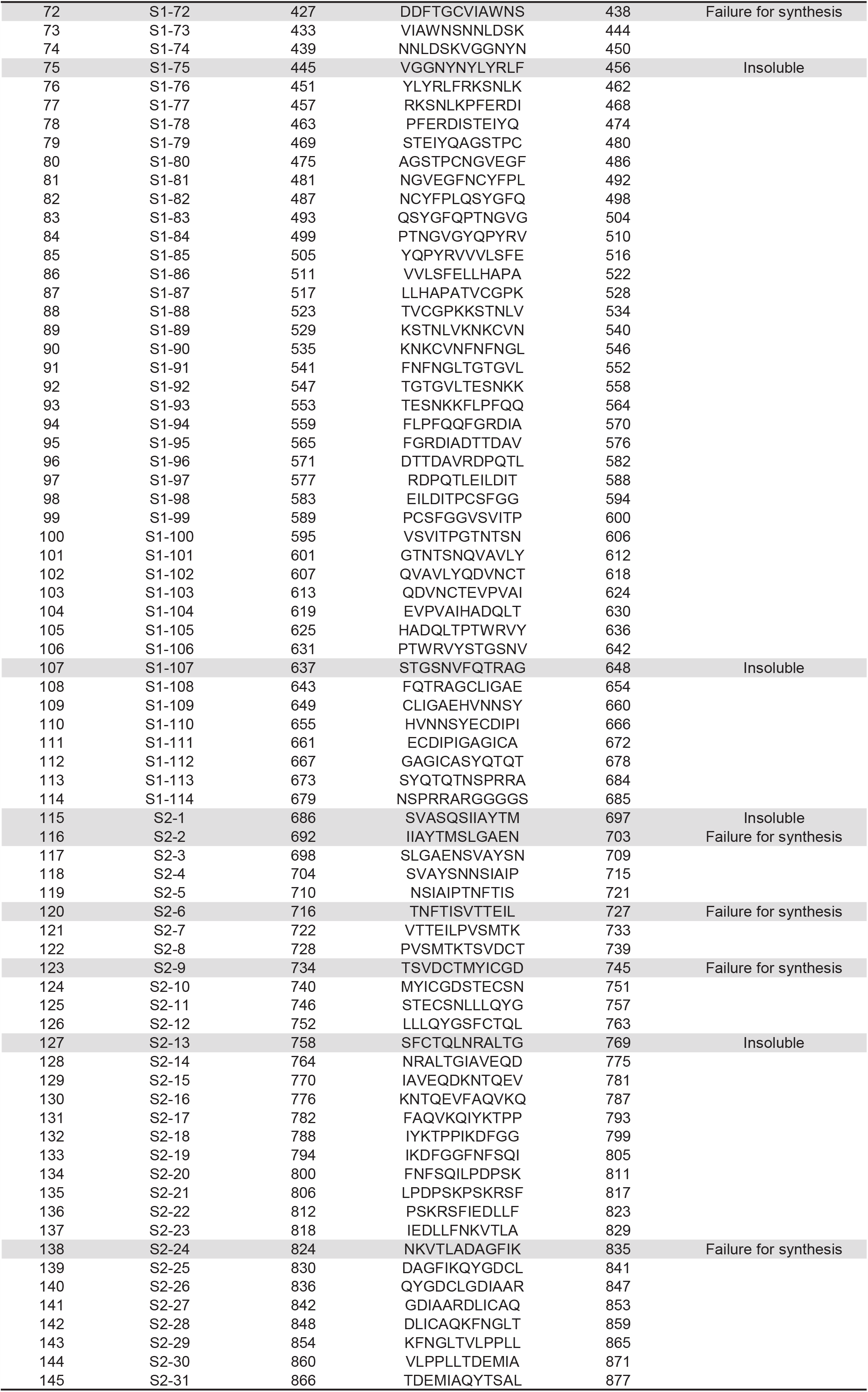
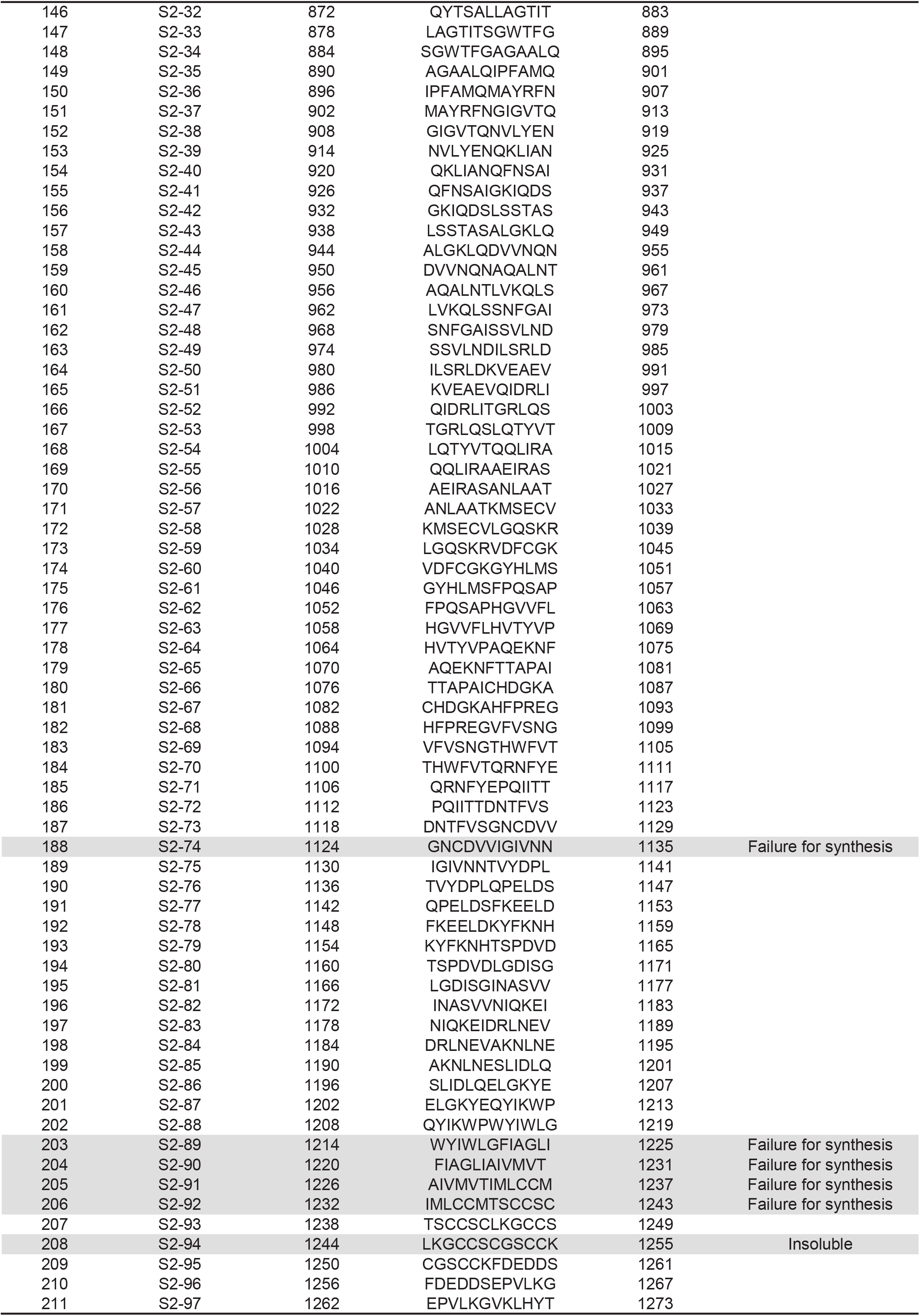
The peptides synthesized in this study.

**Extended Data Table 2.** Serum samples used in this study.

| **SARS-CoV-2 patient group** |  | n=55 |
| --- | --- | --- |
| Gender | Male | 27 |
|  | Female | 28 |
| Age |  | 41.5±14.9 |
| Severity | mild cases | 8 |
|  | moderate cases | 47 |
| Days after onset |  | 27.5±7.7 |
| hospital stay (days) |  | 14.0±5.6 |
| **Non-infection control group** |  | n=18 |
| Lung cancer patients (LC) |  | 9 |
| Health control (HC) |  | 9 |
| Gender | Male | 8 |
|  | Female | 10 |
| Age |  | 50.4±12.5 |
| Sample collection (year) |  | 2017-2018 |

